# Fixel-Based Analysis Identifies Selective Vulnerability of Non-Dominant Fiber Populations in Aging and Cognitive Decline

**DOI:** 10.1101/2025.05.29.25328528

**Authors:** Aditi Sathe, Ankita Chatterjee, Niranjana Shashikumar, Kimberly R. Pechman, Kurt Schilling, Bennett A. Landman, Corey J. Bolton, Panpan Zhang, Andrew R. Bender, Timothy J. Hohman, Angela L. Jefferson, Derek B. Archer

**Affiliations:** Vanderbilt Memory and Alzheimer’s Center, Vanderbilt University School of Medicine, Nashville, TN, USA; Vanderbilt Genetics Institute, Vanderbilt University Medical Center, Nashville, TN, USA; Vanderbilt University Institute of Imaging Science, Vanderbilt University Medical Center, Nashville, TN, USA; Department of Biomedical Engineering, Vanderbilt University, Nashville, TN, USA; Department of Electrical and Computer Engineering, Vanderbilt University, Nashville, TN, US; Department of Medicine, Vanderbilt University Medical Center, Nashville, TN, USA; Department of Biostatistics, Vanderbilt University Medical Center, Nashville, TN; Cleveland Clinic, Lou Ruvo Center for Brain Health, Las Vegas, NV, USA

## Abstract

Fixel-Based Analysis (FBA) offers a novel framework to disentangle fiber-specific white matter (WM) degeneration, particularly within complex crossing-fiber architecture where conventional diffusion tensor imaging (DTI) falls short. In this study, we leveraged single-shell diffusion MRI data from 297 older adults enrolled in the Vanderbilt Memory & Aging Project to quantify fiber density (FD), cross-section (FC), and their composite (FDC) across primary (N1), secondary (N2), and tertiary (N3) fiber populations. By stratifying white matter into anatomically defined crossing-fiber convergence groups, we examined how fiber dominance and architectural complexity modulate associations with age, cognitive status, and longitudinal cognitive decline. Results revealed that FD and FDC in non-dominant (N2/N3) fibers were the strongest predictors of both baseline and longitudinal cognitive trajectories, particularly in memory and executive domains. These associations persisted across fiber convergence strata, suggesting that fiber population identity, rather than anatomical complexity, may confer greater vulnerability to age- and disease-related degeneration. Our findings position non-dominant fiber metrics as sensitive candidate biomarkers for early white matter disruption in Alzheimer’s disease and support the application of multi-fiber modeling to enhance detection of preclinical neurodegeneration.

## 1. Introduction

Age-related white matter (WM) alterations in the brain have been consistently linked to changes in cognitive function in both healthy and impaired aging (Ferro & Madureira, 2002)(Bendlin et al., 2010)(Madden et al., 2009). WM volume (Liu et al., 2017) and tract integrity (Benitez et al., 2018) have been noted to decline with age, particularly through mechanisms that result in myelin damage (Gong et al., 2023) and axonal degeneration (Salvadores et al., 2017). Characterizing the spatial and anatomical specificity of these WM alterations is essential to understanding the biological substrates of cognitive aging (Bartzokis et al., 2004).

Diffusion tensor imaging (DTI) has served as the primary noninvasive modality for assessing WM microstructure (Kennedy & Raz, 2009)(Aung et al., 2013), using scalar metrics such as fractional anisotropy (FA) (Voineskos et al., 2012) and mean diffusivity (MD)(Cox et al., 2016). Accumulating evidence implicates age-related axonal degeneration (Salvadores et al., 2017) and myelin damage (Nasrabady et al., 2018) as critical contributors to the pathophysiological cascade of AD (Bartzokis et al., 2003)(Papuć & Rejdak, 2018), primarily through the selective vulnerability and subsequent degeneration of late-myelinating neurons (Braak et al., 2000). DTI studies consistently demonstrate that the greatest age-related declines in FA occur within thinly myelinated associative WM tracts (Kochunov et al., 2009), whereas heavily myelinated primary motor and sensory pathways exhibit comparatively minimal changes (GarnierLCrussard et al., 2022)(Peter et al., 2025). Notably, disruptions in the microstructural integrity of long-range association fibers, including the superior longitudinal fasciculus (SLF), inferior longitudinal fasciculus (ILF), uncinate fasciculus (UF), and cingulum bundle (Lee et al., 2015)(Huang et al., 2012)(Mayo et al., 2017)(Stone et al., 2021), emerge as some of the most robust and reproducible neuroimaging correlates of cognitive decline across the AD continuum. Recent work from our group supports that these tracts are highly associated with cognitive decline. For example, we found that ILF and the cingulum bundle are associated with cognitive impairment and decline in the Vanderbilt Memory & Aging Project (VMAP), and that microstructure within these tracts has independent contributions to cognitive decline beyond that of hippocampal volume (Archer et al., 2020). Further, large-scale work from our group (n=4,467) has validated that white matter microstructure is intricately linked to cognitive performance and decline across 9 different cohorts (Peter et al., 2025).

A fundamental limitation of the DTI single-tensor model is its inherent inability to resolve complex fiber configurations within a voxel (Landman et al., 2012)(Figley et al., 2022). This is a critical limitation, as regions of fiber crossing, branching, and convergence are estimated to constitute anywhere from 60 to 90% of the brain white matter(Jeurissen et al., 2012). More significantly, previous research has demonstrated that the WM regions most vulnerable to disease-related alterations in cognitive impairment and Alzheimer’s disease (AD), such as the cingulum bundle and SLF, exhibit a high degree of complexity in fiber structure and orientation (Douaud et al., 2011). To identify early microstructural changes that may remain undetected in regions containing only a single fiber population, it is essential to examine complex white matter architecture where affected and unaffected fiber pathways converge (Bender et al., 2025)(Sánchez et al., 2020). An anatomically precise, fiber-specific analytical framework is therefore needed to characterize white-matter vulnerability across the aging spectrum.

Fixel-based analysis (FBA) (D. A. Raffelt et al., 2017) is an emerging approach that uses dMRI data to quantify properties of individual fiber populations or “fixels” within a voxel. FBA enables independent estimation of fiber density (FD), fiber-bundle cross section (FC), and their composite measure — fiber density and cross section (FDC), capturing both microstructural and macrostructural WM integrity on the scale of individual fiber populations (Dhollander et al., 2021). Compared to traditional methods, FBA offers greater anatomical specificity (D. A. Raffelt et al., 2015) and recent applications of FBA have revealed nuanced patterns of WM degeneration in a wide range of neurological disorders (Haykal et al., 2019) (Kirkovski et al., 2020) (Oh et al., 2021) (Finkelstein et al., 2021).

Recent studies have demonstrated that FBA metrics differ significantly, particularly within the fornix(Bhattarai et al., 2023) and SLF (Billaud et al., 2024), across diagnostic groups along the AD continuum (cognitively unimpaired [CU]/mild cognitive impairment [MCI]/dementia due to AD) (Mito et al., 2018) and may also serve as direct correlates of AD-relevant endophenotypes. Specifically, FBA metrics have been shown to exhibit negative correlations with tau-PET uptake (Ahmadi et al., 2024) and positive correlations with memory performance (Billaud et al., 2024). Notably, these associations display substantial regional variability (Tinney et al., 2024), highlighting the heterogeneous nature of microstructural degeneration in early-stage AD (Wen et al., 2019).

Most FBA studies to date have focused exclusively on the dominant (primary) fiber population within each voxel, limiting insight into the vulnerability of non-dominant fiber populations. The present study extends prior work by examining FBA metrics across primary (N1), secondary (N2), and tertiary (N3) fiber populations throughout the brain. We evaluated global associations of FD, FC, and FDC with age, cognitive status (CU vs. MCI), and baseline and longitudinal cognitive performance. To further examine how complex fiber architecture modulates WM degeneration, we stratified brain regions into low, medium, high, and maximum crossing-fiber groups based on the number of distinct fiber populations within a voxel. To our knowledge, this is the first study to stratify white matter regions by in vivo crossing-fiber concentration to examine how anatomical fiber complexity modulates age-related and pathological white matter degeneration across multiple fiber populations.

We hypothesized that secondary and tertiary fiber populations may subserve distinct functional pathways and may be differentially susceptible to aging-related or pathological degeneration due to their lower structural dominance and spatial orientation within dense crossing fiber regions. Accordingly, we expected these fiber populations to demonstrate stronger associations with age, diagnostic differences, and cognitive decline than primary fibers. We further hypothesized that regions with the highest convergence of crossing-fiber would exhibit the most pronounced reductions across all FBA metrics.

Given the known role of crossing fiber regions in maintaining large-scale brain network efficiency (Wiegell et al., 2000)(Hua et al., 2009), disruption within these areas may contribute disproportionately to cognitive decline in aging and AD.

## 2. Methods

### 2.1 Study Cohort

The Vanderbilt Memory & Aging Project (VMAP) (Jefferson et al., 2016) is a longitudinal observational study investigating vascular health and brain aging, including participants aged 60+ years who are considered CU or have MCI. MCI determinations were based upon National Institute on Aging/Alzheimer’s Association Workgroup core clinical criteria (Albert et al., 2011). VMAP participants (n_participants_=297, n_observations_=1256, age at baseline: 73 ± 7 years, 44% MCI) underwent longitudinal neuropsychological assessment and brain MRI over a maximum of six time points including baseline (n=325), 18-month (n=292), 3-year (n=274), 5-year (n=233), 7-year (n=158), and 9-year (n=93) visits.

The mean follow-up period was 4.4 ± 2.5 years. At study entry, participants completed a comprehensive evaluation, including fasting blood draw, physical examination, clinical interview with medication review, echocardiogram, and brain MRI (Moore et al., 2021). As part of the extensive screening, participants were excluded for a cognitive diagnosis of dementia, magnetic resonance imaging (MRI) contraindication, history of neurological disease (e.g., multiple sclerosis, stroke), heart failure, major psychiatric illness, head injury with loss of consciousness >5 minutes, or a systemic or terminal illness affecting follow-up participation. Informed consent was provided by all participants and the Vanderbilt Institutional Review Board approved the protocol. Several demographic and clinical covariates were required for inclusion in the present study, including age, sex, educational attainment, race/ethnicity, apolipoprotein E (*APOE*) carrier status (ε2, ε3, ε4), Framingham Stroke Risk Profile (FSRP; excluding points for age), and cognitive status at baseline (CU, MCI).

### 2.2 Neuropsychological Assessment

Participants completed a common, comprehensive neuropsychological protocol assessing language, information processing speed, executive function, visuospatial skills, and episodic learning and memory. As previously described (Kresge et al., 2018), we created psychometrically sound composite measures for memory and executive function. The memory composite compiled scores from the California Verbal Learning Test-Second Edition (CVLT-II) Total Learning, Interference Condition, Long Delay Free Recall, and Recognition components, together with identical components of the Biber Figure Learning Test (Gifford et al., 2020; Glosser et al., 2002). For the executive functioning composite, the Delis-Kaplan Executive Function System (DKEFS) Tower Test, DKEFS Letter-Number Switching, DKEFS Color-Word Inhibition, and Letter Fluency (FAS) test scores were evaluated. The Animal Naming Test and Boston Naming Test scores were used to quantify language performance, and WAIS-IV Coding, and DKEFS Number Sequencing scores assessed information processing speed. The Hooper Visual Organization Test was used to evaluate visuospatial skills.

### 2.3 Diffusion MRI Acquisition and Preprocessing

Participants were scanned at the Vanderbilt University Institute of Imaging Science on a 3 T Philips Achieva system (Best, The Netherlands). Imaging parameters have been previously outlined (Jefferson et al., 2016; Moore et al., 2018). Briefly, dMRI images with a spin-echo echo-planar sequence, repetition time = 10 s, echo time = 60 ms, spatial resolution = 2mm isotropic, and b-values: 0, 1000 s/mm2 were acquired. dMRI images were collected along 32 diffusion gradient vectors together with one non-diffusion (B0) weighted image. The PreQual (Cai et al., 2021) pipeline was used to preprocess all dMRI data, ensuring robust preprocessing (e.g., denoising, eddy current correction, motion correction). The quality control PDFs generated by the PreQual pipeline underwent manual inspection, and participants with poor data quality were excluded from the analysis. Generally, imaging sessions were removed due to inaccurate synthetic b0 creation, inaccurate brain masking, and excessive motion.

### 2.4 Quantifying Fixel Metrics

Fixel-based metrics were computed using the MRtrix3 software suite, following standard recommended procedures. Diffusion-weighted imaging (dMRI) data were resampled to an isotropic voxel size of 1.25 mm³. Response functions were estimated using the Tournier algorithm and averaged across all participants to derive a group-representative response function. Although the dataset was single-shell, multi-shell multi-tissue constrained spherical deconvolution (MSMT-CSD) was employed to compute fiber orientation distributions (FODs), in order to take advantage of the algorithm’s hard non-negativity constraint (Newman et al., 2020)(Jeurissen et al., 2014). To enable spatial correspondence across subjects, a study-specific FOD population template was generated from a representative subset of 30 participants, including 15 individuals from each of the CU and MCI groups. All individual FOD images were non-linearly registered to this population template, and fixel-based metrics were subsequently derived for each subject. **Figure 1** shows representative grids from FOD, fixel, and primary fiber population (N1) fiber density maps.

**Figure 1:**
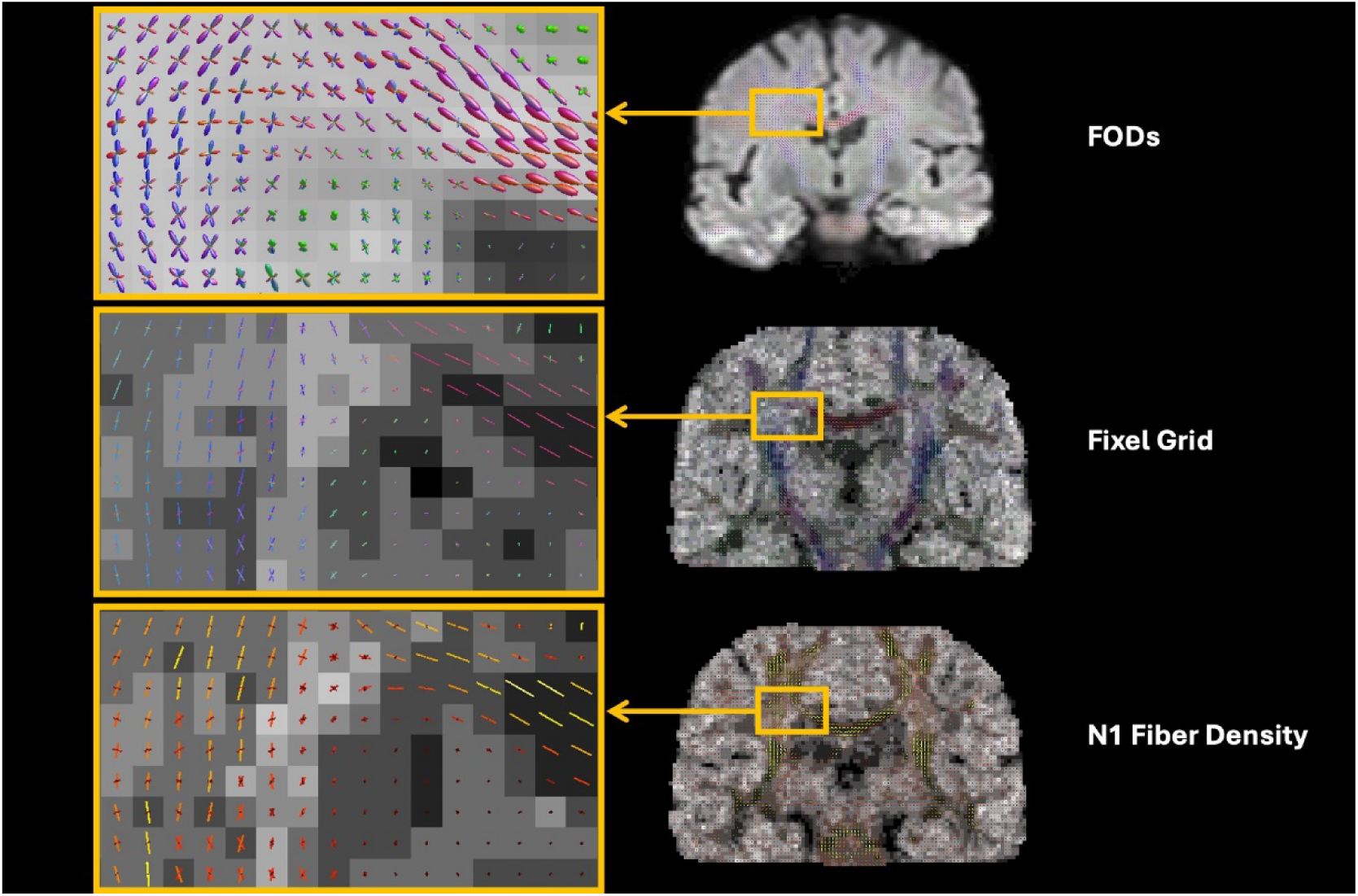
Computation of Fixel-wise Apparent Fiber Density from White Matter FODs. Top: White matter fibre orientation distributions (WM FODs), estimated using three-tissue constrained spherical deconvolution (CSD), serve as the basis for both the fixel grid and the apparent FD metric. FODs are visualized in a coronal slice overlaid on an FOD-based directionally encoded colour (DEC) map (red = mediolateral, green = anteroposterior, blue = superoinferior).Middle: Fixels are derived by segmenting each voxel’s FOD into discrete lobes corresponding to distinct fibre populations, based on local angular maxima. In contrast to the regular voxel lattice, the spatial distribution and orientation of fixels are anatomically constrained by local white matter structure. Bottom: Apparent fibre density is calculated for each fixel as the integral of its corresponding FOD lobe. Voxel-wise FD (underlying grayscale intensities) reflects the sum of all fixel-wise FD values within each voxel. All FD values are presented in arbitrary units.

The following fixel metrics were computed and analyzed (illustrated in **Figure 2**):

- Fiber Density (FD): A subvoxel-level metric reflecting the integral of the FOD along a given fixel direction. FD serves as an estimate of intra-axonal volume within that fiber population.
- Fiber-Bundle Cross-Section (FC): A macrostructural measure quantifying the morphological differences in fiber-bundle diameter between an individual subject and the population template. It is computed as the degree of deformation required in the plane perpendicular to the fixel orientation during spatial registration. All FC values reported in this study refer to the logarithmic transform of FC (log-FC).
- Fiber Density and Cross-Section (FDC): A combined metric that integrates both FD and FC, enabling simultaneous assessment of microstructural and macrostructural white matter properties.

**Figure 2:**
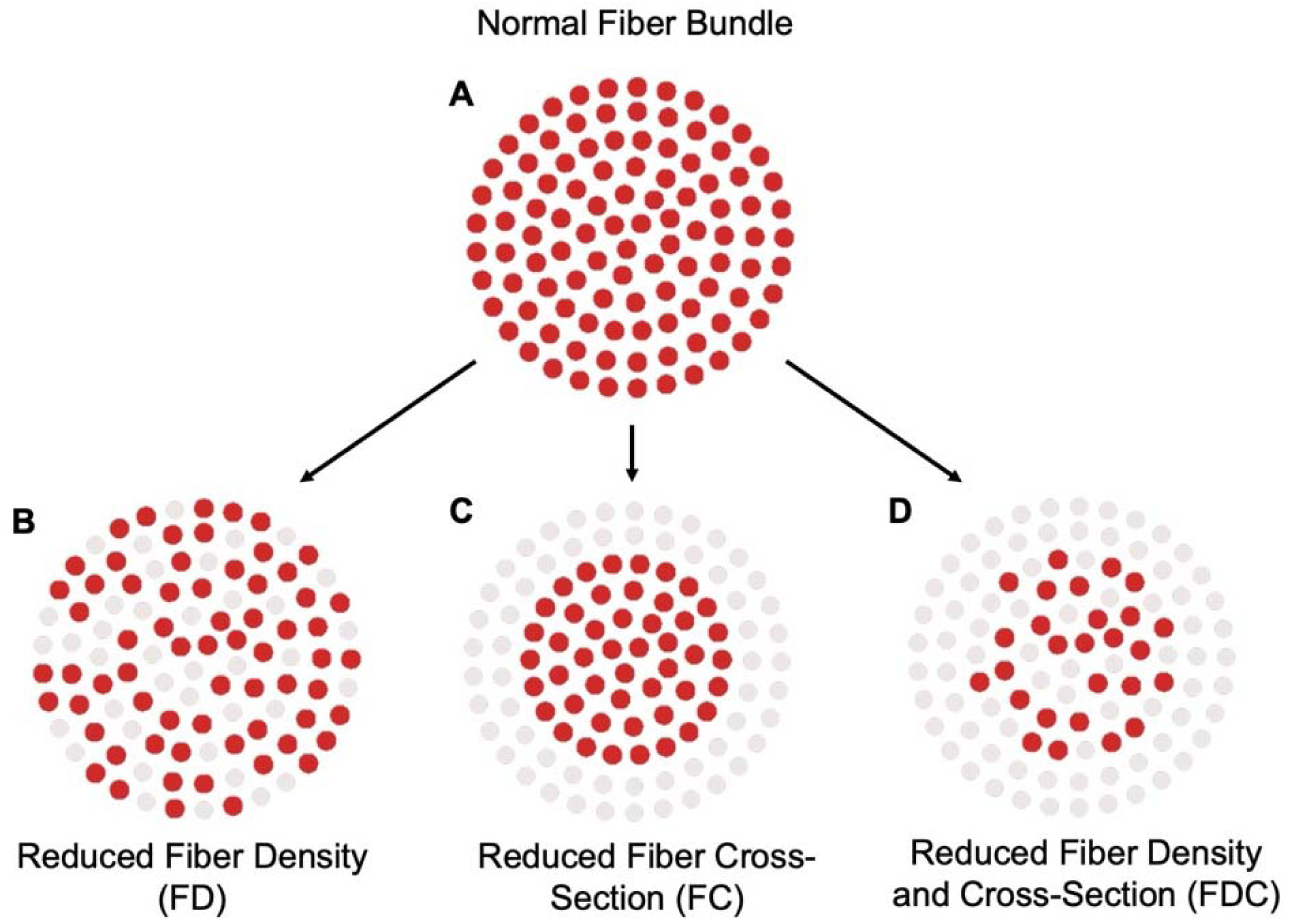
Illustration of FBA metrics, consistent with the original schematic presented in Raffelt et al. (2017). FBA decomposes intra-axonal volume into three components: FD (microstructure), FC (macrostructure), and FDC (combined). A: Normal fiber architecture. B: Isolated FD reduction indicating axonal loss. C: FC reduction reflecting fiber-bundle contraction relative to the population template. D: Joint reduction in FD and FC.

### 2.5 ROI Segmentation Using a Crossing Fiber Atlas

ROI segmentation into low, medium, high, and maximum crossing-fiber groups was achieved using the TractSeg voxel-wise bundle overlap map (Schilling et al., 2021). The TractSeg pipeline (https://github.com/MIC-DKFZ/TractSeg) was applied to each subject’s FOD data to generate 72 anatomically defined white matter bundles. Resulting bundle-specific tractograms were transformed nonlinearly into the study-specific FBA template space, where streamlines were assigned to their corresponding fixels and voxels. Within template space, the number of unique bundles associated with each fixel was quantified and aggregated to yield a voxel-wise bundle count.

In the current study, voxels containing one or fewer bundles (below the 32.5^th^ percentile) were excluded from ROI analysis to focus on regions with true crossing-fiber architecture. We leveraged the population-average bundle count map and defined crossing-fiber groups by percentile intensity thresholds as follows:

- Low (32.5–50th percentile)
- Medium (50–75th percentile)
- High (75–87.5th percentile)
- Maximum (87.5–100th percentile)

Each group’s segmented map served as a weighted ROI mask co-registered to the FBA template. For each subject, FD, FC, and FDC values were averaged within these weighted masks to quantify FBA metrics for each crossing-fiber group. **Figure 3** displays representative cross-sections of the four ROIs in template space.

**Figure 3:**
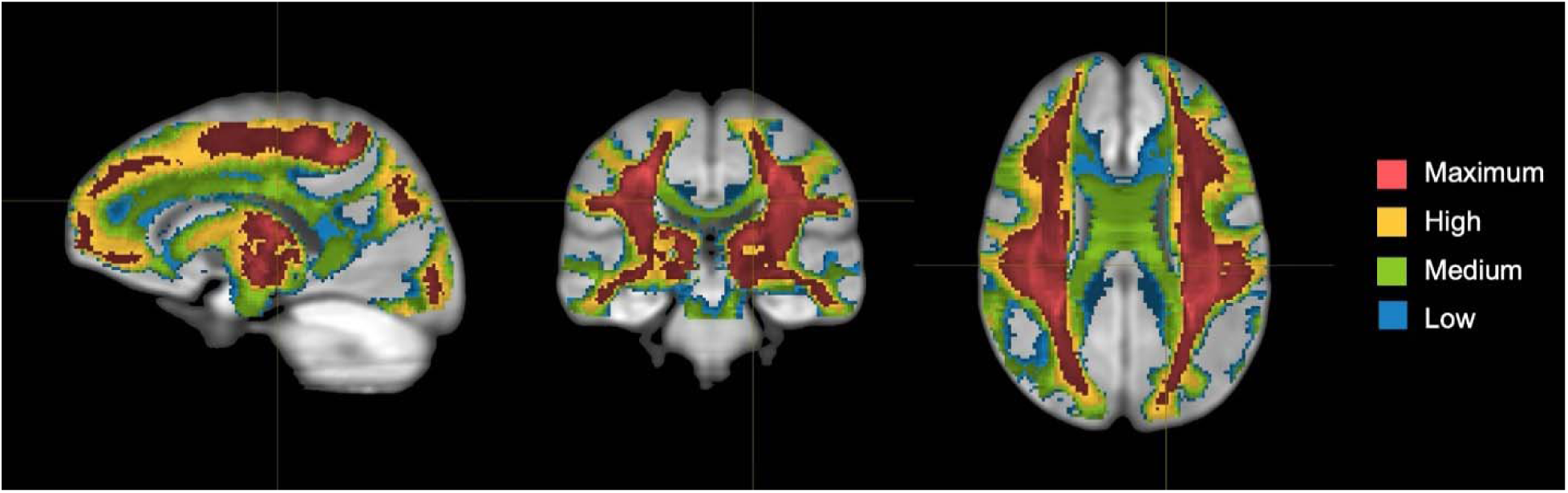
Sagittal, coronal, and axial views of crossing fiber convergence groups. White matter ROIs are stratified by crossing fiber convergence based on diffusion-derived metrics from Schilling et al. (2021). Color-coded overlays indicate voxel-wise fiber complexity: blue (low, absolute range 1–2.14; 32.5–50th percentile of robust range), green (medium, 2.14–4.16; 50–75th), yellow (high, 4.16–6.00; 75–87.5th), and red (maximum, 6.00–16.26; 87.5–100th). Segmentations delineate major tract crossing zones, reflecting regions of increased fiber intersection. Images are shown in neurological orientation with standardized intensity scaling.

### 2.6 Statistical Analyses

All statistical analyses were performed in R version 4.4.2 (2024-10-31) (http://www.r-project.org/). Covariates included age, sex, cognitive status (CU, MCI), race/ethnicity, FSRP scores (excluding points for age), and APOE-ε4 carrier status. *APOE*-ε4 carrier status was defined as positive (ε2/ε4, ε3/ ε4, ε4/ ε4) or negative (ε2/ε2, ε2/ε3, ε3/ε3). All predictors were standardized before analyses, and an FDR procedure was used to correct for multiple comparisons. We first conducted independent samples *t*-tests to assess group differences between CU and MCI participants across FBA metrics (FD, FC, FDC) in each of the top three most prominent fiber populations (N1, N2, N3). We then used linear regression models to estimate the association between each FBA metric and age across the entire sample.

Baseline effects of FBA-derived metrics on cognitive performance were evaluated using general linear models. We also examined interactions between FBA metrics and diagnostic group in predicting baseline cognitive performance. To assess the relationship between FBA metrics and longitudinal cognitive change, we employed linear mixed-effects regression models with random intercepts to model the association between baseline FBA metrics and cognitive trajectories over time. We further tested the interaction between each FBA metric and diagnosis in predicting longitudinal change in cognition.

All analyses described above were repeated separately within each crossing-fiber convergence group. Additionally, to examine differences across anatomical fiber complexity, we performed pairwise *t*-tests between crossing-fiber groups (e.g., low vs medium, medium vs high, high vs max) for each FBA metric and fiber population.

## 3. Results

### 3.1 Participant and Crossing-Fiber Group Characteristics

Significant group differences were observed between CU and MCI participants in years of education (p < .01), *APOE*-ε4 carrier status (p < .02), and all baseline cognitive scores, including memory composite, executive function composite, language, information processing speed, and visuospatial skills (all p < .001). Full demographic and cognitive data are provided in **Table 1**.

**Table 1:**
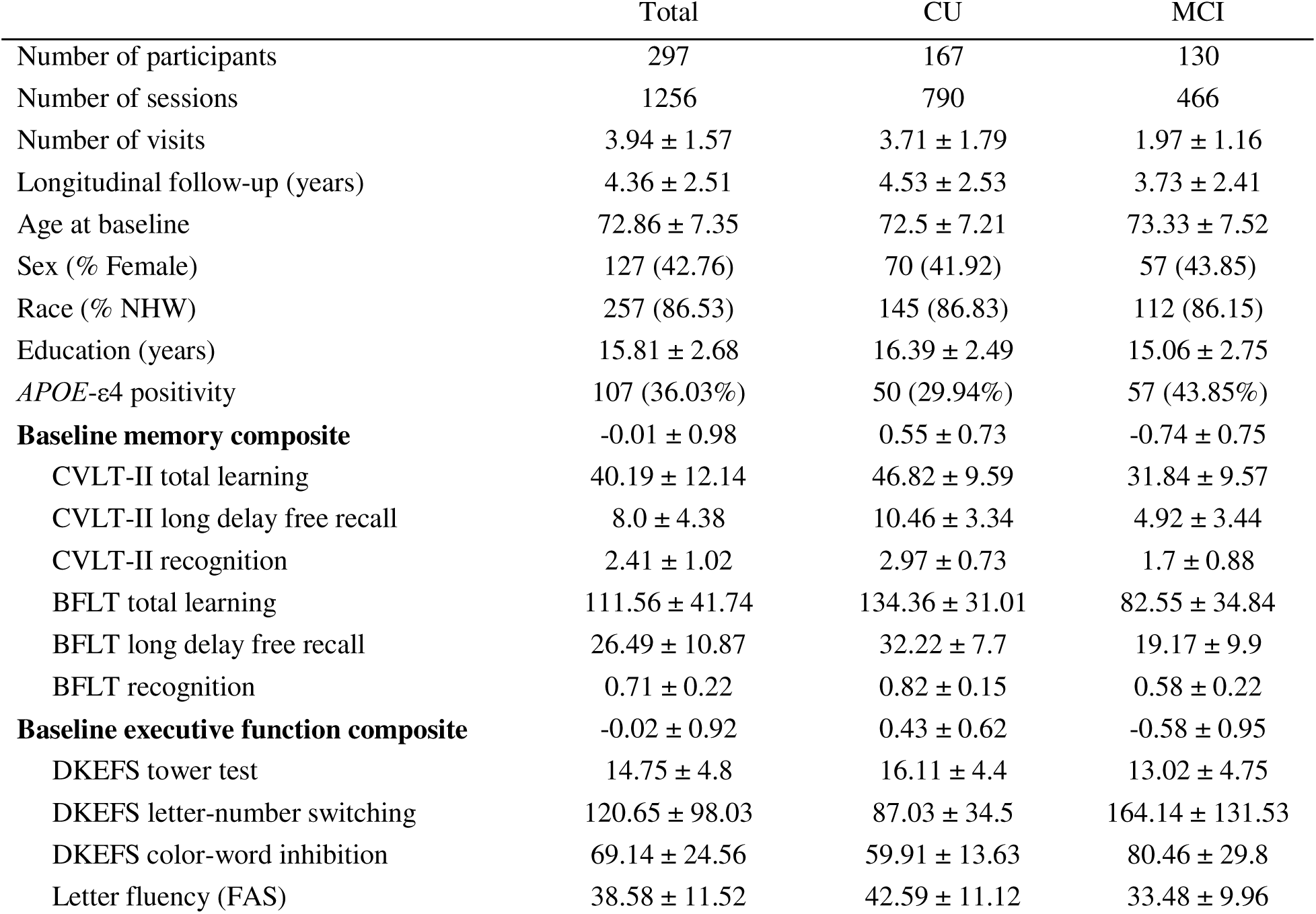
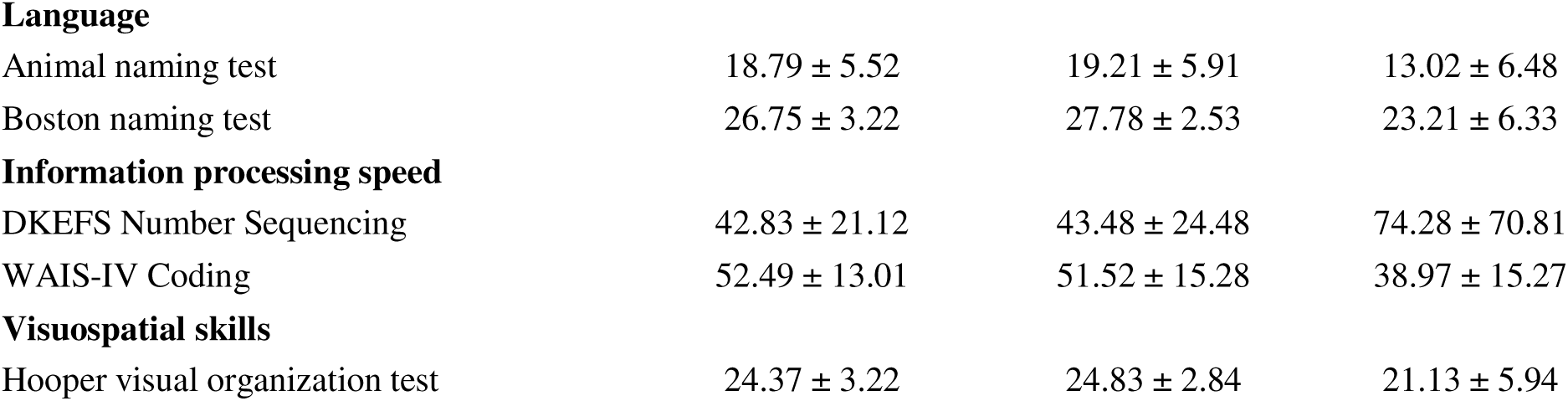
Vanderbilt Memory and Aging Project (VMAP) cohort information.

|  | Total | CU | MCI |
| --- | --- | --- | --- |
| Number of participants | 297 | 167 | 130 |
| Number of sessions | 1256 | 790 | 466 |
| Number of visits | $3.94 \pm 1.57$ | $3.71 \pm 1.79$ | $1.97 \pm 1.16$ |
| Longitudinal follow-up (years) | $4.36 \pm 2.51$ | $4.53 \pm 2.53$ | $3.73 \pm 2.41$ |
| Age at baseline | $72.86 \pm 7.35$ | $72.5 \pm 7.21$ | $73.33 \pm 7.52$ |
| Sex (% Female) | 127 (42.76) | 70 (41.92) | 57 (43.85) |
| Race (% NHW) | 257 (86.53) | 145 (86.83) | 112 (86.15) |
| Education (years) | $15.81 \pm 2.68$ | $16.39 \pm 2.49$ | $15.06 \pm 2.75$ |
| *APOE*-$\epsilon 4$ positivity | 107 (36.03%) | 50 (29.94%) | 57 (43.85%) |
| **Baseline memory composite** | $-0.01 \pm 0.98$ | $0.55 \pm 0.73$ | $-0.74 \pm 0.75$ |
| CVLT-II total learning | $40.19 \pm 12.14$ | $46.82 \pm 9.59$ | $31.84 \pm 9.57$ |
| CVLT-II long delay free recall | $8.0 \pm 4.38$ | $10.46 \pm 3.34$ | $4.92 \pm 3.44$ |
| CVLT-II recognition | $2.41 \pm 1.02$ | $2.97 \pm 0.73$ | $1.7 \pm 0.88$ |
| BFLT total learning | $111.56 \pm 41.74$ | $134.36 \pm 31.01$ | $82.55 \pm 34.84$ |
| BFLT long delay free recall | $26.49 \pm 10.87$ | $32.22 \pm 7.7$ | $19.17 \pm 9.9$ |
| BFLT recognition | $0.71 \pm 0.22$ | $0.82 \pm 0.15$ | $0.58 \pm 0.22$ |
| **Baseline executive function composite** | $-0.02 \pm 0.92$ | $0.43 \pm 0.62$ | $-0.58 \pm 0.95$ |
| DKEFS tower test | $14.75 \pm 4.8$ | $16.11 \pm 4.4$ | $13.02 \pm 4.75$ |
| DKEFS letter-number switching | $120.65 \pm 98.03$ | $87.03 \pm 34.5$ | $164.14 \pm 131.53$ |
| DKEFS color-word inhibition | $69.14 \pm 24.56$ | $59.91 \pm 13.63$ | $80.46 \pm 29.8$ |
| Letter fluency (FAS) | $38.58 \pm 11.52$ | $42.59 \pm 11.12$ | $33.48 \pm 9.96$ |
| Language |  |  |  |
| Animal naming test | $18.79 \pm 5.52$ | $19.21 \pm 5.91$ | $13.02 \pm 6.48$ |
| Boston naming test | $26.75 \pm 3.22$ | $27.78 \pm 2.53$ | $23.21 \pm 6.33$ |
| Information processing speed |  |  |  |
| DKEFS Number Sequencing | $42.83 \pm 21.12$ | $43.48 \pm 24.48$ | $74.28 \pm 70.81$ |
| WAIS-IV Coding | $52.49 \pm 13.01$ | $51.52 \pm 15.28$ | $38.97 \pm 15.27$ |
| Visuospatial skills |  |  |  |
| Hooper visual organization test | $24.37 \pm 3.22$ | $24.83 \pm 2.84$ | $21.13 \pm 5.94$ |

Pairwise t-tests between crossing-fiber groups (e.g., low vs. medium, medium vs. high, high vs. max) revealed significant differences in the values of nearly all fixel-based metrics (FD, FC, FDC) for each bin comparison across fiber population levels (all p < .001). Metrics were derived separately for primary (N1), secondary (N2), and tertiary (N3) fiber populations. Complete bin-wise statistics are reported in **Supplementary Table 1**.

Between-group comparisons (CU vs. MCI) for all FBA metrics showed significant group differences across bins and fiber populations. The strongest effects were observed for FD of the tertiary fiber population (N3) in the Maximum and High crossing fiber groups, respectively. Overall, MCI participants exhibited lower FD, FC, and FDC compared to CU individuals.

Age was significantly associated with all global FBA-derived metrics, including FD, FC, and FDC across N1, N2, and N3 fiber populations (all p < .001). N3 FD yielded both the lowest p-value and the highest adjusted R² among all metrics. **Figure 4** illustrates the associations between age and scaled fixel metrics across N1, N2, and N3 fiber populations. Region-stratified analyses by crossing-fiber density group (low, medium, high, maximum) mirrored these findings. The lowest p-values and highest adjusted R² values were observed for N3 FD in the low, medium, and high bins, respectively. Full model statistics are provided in **Supplementary Table 2**.

**Figure 4:**
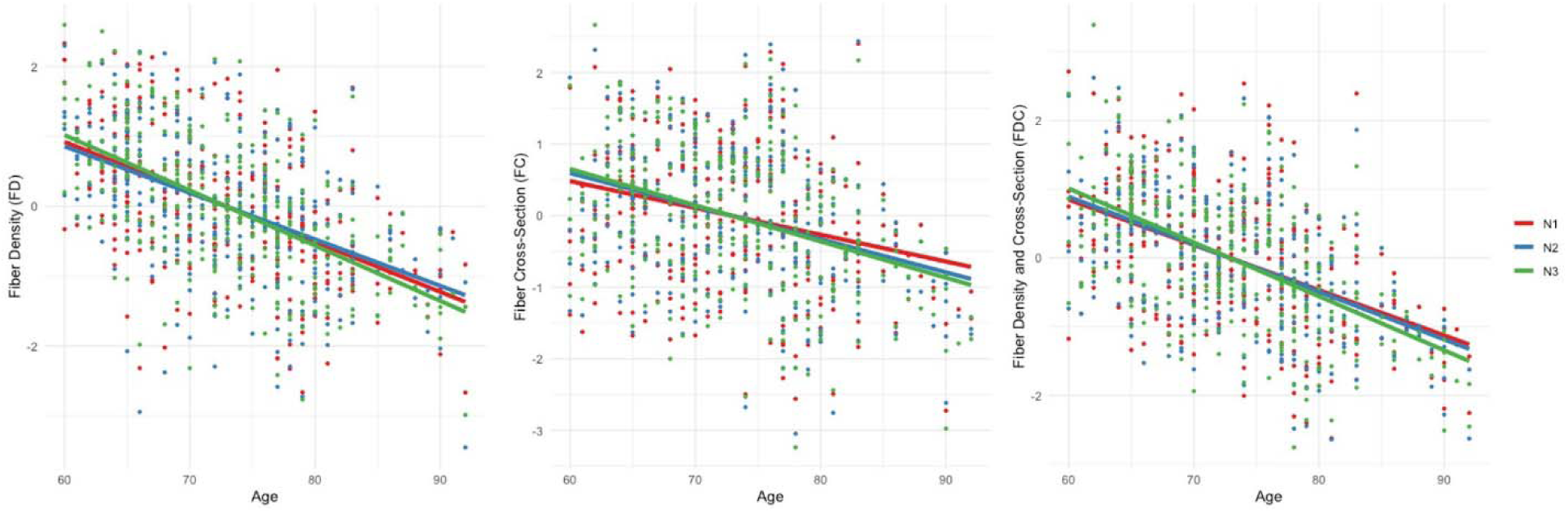
Associations between age and scaled fixel-based metrics across fiber populations. Scatterplots with fitted regression lines illustrate the relationship between age (x-axis) and standardized values (z-scored) of fiber density (FD), fiber cross-section (FC), and fiber density and cross-section (FDC) on the y-axes. Each panel represents one FBA metric. Individual data points and regression lines are color-coded by fiber population: primary (red), secondary (blue), and tertiary (green).

### 3.3 FBA Associations with Baseline Cognition

Global models examining baseline cognitive scores showed significant associations between fixel-based metrics and performance. For memory, N3 FD and N2 FD were significantly associated (p < .05). For executive function, N3 FDC and N2 FDC were the top predictors (p < .01). The top three predictors with the lowest p-value and the highest adjusted R² for memory and executive function composite scores are presented in **Table 2**. Full cognitive associations across all FBA metrics and cognitive domains are provided in **Supplementary Table 3**. We did not observe any FBA metric *×* diagnosis interactions on cognitive scores at baseline. Results for this interaction analysis can be found in **Supplementary Table 4**.

**Table 2:** Global FBA metrics associations with baseline cognition.

| Outcome | Predictor | $\beta$ | $\beta_{SE}$ | t | p | $p_{FDR}$ | Adjusted R2 |
| --- | --- | --- | --- | --- | --- | --- | --- |
| Composite memory score |  |  |  |  |  |  |  |
|  | N3 FD | 1.92 | 0.74 | 2.59 | 1.01E-02 | 2.63E-02 | 53.55% |
|  | N2 FD | 2.13 | 0.83 | 2.56 | 1.10E-02 | 2.76E-02 | 53.53% |
|  | N1 FD | 1.62 | 0.79 | 2.06 | 4.01E-02 | 8.77E-02 | 53.16% |
| Composite executive function score |  |  |  |  |  |  |  |
|  | N2 FDC | 1.51 | 0.43 | 3.54 | 4.63E-04 | 4.01E-03 | 47.82% |
|  | N3 FDC | 1.46 | 0.42 | 3.46 | 6.12E-04 | 4.01E-03 | 47.73% |
|  | N2 FC | 0.20 | 0.06 | 3.46 | 6.30E-04 | 4.01E-03 | 47.72% |

Analyses stratified by crossing-fiber group revealed similar trends, with N3 FD and N2 FD showing significant associations with memory, and N3 FDC and N2 FDC with executive function (all p < .05). The only exception was observed in the maximum crossing-fiber group, where none of the FBA metrics reached significance for associations with memory. Complete group-wise results are available in **Supplementary Table 5**.

### 3.4 FBA Associations with Longitudinal Cognition

In models predicting longitudinal change in cognition, a larger number of fixel-based metrics reached statistical significance. For memory, N3 FD and N2 FD remained the most significant predictors across all models (p < .01). For executive function, N3 FDC and N2 FDC yielded the strongest effects (p < .001). **Table 3** lists the three FBA metrics yielding the lowest p-values for memory and executive function decline. Detailed longitudinal associations for all global FBA metrics and cognitive domains are available in **Supplementary Table 6**. We also evaluated FBA metric *×* diagnosis interactions on longitudinal cognitive scores. Results for these analyses can be found in **Supplementary Table 7**. Most FBA metrics showed significant interaction with diagnosis, with the lowest FDR-corrected p-values achieved by N3 FD for memory and N3 FC for executive function.

**Table 3:** Global fixel metrics associations with longitudinal cognition.

| Outcome | Predictor | $\beta$ | $\beta_{SE}$ | t | p | $p_{FDR}$ |
| --- | --- | --- | --- | --- | --- | --- |
| Composite memory score |  |  |  |  |  |  |
|  | N3 FD | 2.86 | 0.75 | 3.81 | 1.38E-04 | 5.69E-04 |
|  | N2 FD | 2.89 | 0.85 | 3.42 | 6.27E-04 | 1.76E-03 |
|  | N1 FD | 2.37 | 0.80 | 2.98 | 2.92E-03 | 7.05E-03 |
| Composite executive function score |  |  |  |  |  |  |
|  | N3 FDC | 1.89 | 0.42 | 4.49 | 7.1293E-06 | 5.55E-05 |
|  | N2 FDC | 1.91 | 0.43 | 4.46 | 8.3463E-06 | 5.84E-05 |
|  | N2 FC | 0.26 | 0.06 | 4.34 | 1.4242E-05 | 9.06E-05 |

When stratified by crossing-fiber group, consistent trends were observed across the low, medium, and high bins. In these groups, N3 FD and N2 FD were the top predictors of memory decline, and N3 FDC and N2 FDC best predicted executive function decline. In the maximum crossing-fiber density group, predictor rankings diverged and showed reduced consistency across fiber populations. Here, the top predictors were N3 FD and N3 FDC for memory decline, and N2 FC and N3 FDC for executive function decline. Full stratified longitudinal results are included in **Supplementary Table 8**.

## 3. Discussion

This study demonstrates that FBA detects robust age- and disease-related alterations in crossing fiber regions across the whole brain WM that are not readily captured by conventional DTI. By quantifying FD, FC, and FDC across primary, secondary, and tertiary fiber populations, we provide evidence that non-primary fiber populations (i.e., N2 and N3 fixels) exhibit stronger associations with cognitive performance and decline in aging populations. These results suggest that tertiary and secondary fiber metrics may offer more sensitive markers of early cognitive vulnerability than single-fiber analyses. FD in N3 and N2 was the strongest predictor of memory decline while FDC in these fiber populations best explained changes in executive function. These findings suggest that memory impairment is primarily linked to microstructural degeneration, whereas executive dysfunction involves both micro- and macrostructural damage. The shared involvement of non-primary fibers highlights the dependence of these cognitive domains on complex distributed structural networks. Our findings agree with prior work from our group which found that the limbic tracts were most associated with memory performance, but a more widespread network of white matter tracts were associated with executive function (Peter et al., 2025).

Although stratified analyses were conducted to evaluate the role of anatomical fiber convergence, we did not find consistent evidence that regions with higher crossing-fiber convergence exhibited more pronounced degeneration in any fiber population. The results largely mirrored global effects, with no substantial differences in significance or effect size across low, medium, high, and maximum convergence groups. These findings suggest that fiber population (primary vs. non-primary) may be a more salient determinant of vulnerability than anatomical convergence alone. It is possible that degeneration occurs diffusely across the white matter regardless of local crossing complexity in early stages of aging and MCI, or that the grouping approach lacked sufficient resolution to capture spatial differences. Our grouping method was conducted without considering lobular- or tract-specific differences. Additional work parcellating crossing-fibers in a more spatially specific manner may provide additional insight into fiber architecture and vulnerability. Alternatively, high fiber convergence may confer redundancy or resilience, rather than vulnerability, under some conditions.

The distinct patterns observed across FBA metrics are consistent with their underlying biological interpretations. FD reflects intra-axonal volume and axonal packing density, FC captures morphological atrophy of fiber bundles, and FDC represents a composite of micro- and macrostructural integrity(D. A. Raffelt et al., 2017). The stronger associations observed with FD and FDC relative to FC suggest early involvement of axonal microstructure, particularly in non-dominant fibers that may subserve distinct functional pathways. These likely represent thinly myelinated tracts, which are especially susceptible to metabolic stress and degeneration. Their selective vulnerability supports the “last in, first out” model of brain aging, wherein later-myelinating pathways show accelerated decline with advancing age (Fjell et al., 2014)(Brickman et al., 2012). This interpretation is supported by DTI studies, such as those by Kochunov et al., which demonstrate that age-related degradation of thinly myelinated tracts parallels reductions in cerebral glucose metabolism. Similarly, multimodal imaging has shown that WM integrity is positively associated with both FDG-PET uptake (Kuczynski et al., 2010) and levels of N-acetyl aspartate (Charlton et al., 2006), a marker of axonal health (Moffett et al., 2007). Together, these findings support a model of progressive white matter degeneration driven by axonal loss, consistent with the “disconnection” hypothesis of cognitive aging, which attributes cognitive decline to impaired network integration arising from structural disintegration(O’Sullivan et al., 2001). It is important to note that FD approximates the total intra-axonal volume within a voxel along a given fixel but cannot differentiate between variations in axon number and diameter, as both contribute to the metric (Dhollander et al., 2021). At lower diffusion weightings, such as b = 1000 s/mm² used in this study, extra-axonal signal contamination further compromises the specificity of FD, limiting its biological interpretability (D. Raffelt et al., 2012). Additionally, FBA is largely insensitive to myelin, as myelin-associated water has a short T2 relaxation time and contributes minimally to the diffusion MRI signal (Dhollander et al., 2021).

The study has several methodological strengths. The use of FBA allowed fiber-specific resolution in regions of complex fiber architecture, overcoming key limitations of traditional DTI metrics. This study sets up a framework that can be used to examine WM integrity in existing older datasets and long-running cohort studies that have standardized dMRI protocols. By modeling primary, secondary, and tertiary fiber populations separately, the analysis captured a granularity of degeneration patterns not previously addressed in most aging or AD studies. The inclusion of longitudinal cognitive outcomes added predictive value to the analysis and enhances the translational potential of the findings. Additionally, the stratification of analyses by crossing-fiber density permitted a more anatomically informed approach, contextualizing microstructural changes within the spatial heterogeneity of white matter architecture.

Nevertheless, several limitations should be acknowledged. The use of dMRI data with a b-value lower than the recommended threshold for constrained spherical deconvolution may have insufficient specificity for proper quantitative interpretation; however, we must note that b≥1,000 s/mm2 and 32+ gradient directions may be sufficient for FBA workflows (Dhollander et al., 2021). While the pipeline applied is validated for single-shell datasets(Luo et al., 2021)(Petersen et al., 2022)(Nguyen et al., 2021), future studies using high b-value HARDI data are likely to improve both accuracy and sensitivity. The imaging data were cross-sectional, limiting conclusions about within-subject progression of white matter degeneration; future work should incorporate longitudinal FBA to capture temporal trajectories. Analyses were conducted using global and group-averaged metrics rather than spatially resolved voxel-wise or tract-specific approaches. Although this design reduced the risk of multiple comparisons, it precludes precise localization of affected tracts. Finally, potential confounding variables such as physical activity and sleep quality were not explicitly modeled and may have influenced the observed associations.

Our findings highlight the promise of FBA, particularly FD and FDC in secondary and tertiary fibers, as candidate biomarkers for early detection and progression of white matter degeneration. By resolving fiber-specific white matter properties, FBA offers a framework for elucidating the microstructural basis of cognitive decline. Future research should investigate how FBA metrics integrate with other biomarkers of AD pathology (e.g., amyloid/tau PET, gray matter atrophy, plasma-based biomarkers). Additionally, validation in independent, multi-site cohorts using harmonized imaging protocols is necessary to establish reproducibility. Studies in genetically at-risk individuals and prodromal stages of neurodegeneration will be particularly informative for determining the earliest manifestations of fiber-specific vulnerability. Finally, FBA may hold promise as a monitoring tool in clinical trials targeting white matter preservation or repair, and its utility in these contexts should be explored in prospective studies.

## 4. Conclusion

This study demonstrates that FBA offers a sensitive and fiber-specific approach for characterizing WM alterations associated with aging and MCI. By quantifying fiber density, cross section, and their combination across primary, secondary, and tertiary fiber populations, we identified distinct patterns of degeneration not captured by traditional DTI methods. The consistent predictive value of non-dominant fiber metrics for cognitive decline underscores their potential clinical utility. Importantly, the lack of strong evidence for selective vulnerability in high-density crossing regions suggests that fiber population identity may be a more critical factor than anatomical convergence in early white matter degeneration. These findings advance our understanding of the microstructural complexity of white matter aging and highlight the value of multi-fiber modeling for capturing subtle changes in brain connectivity. The methodological framework applied here establishes a foundation for future longitudinal and multi-modal studies aiming to refine early biomarkers of neurodegeneration and improve the anatomical specificity of neuroimaging tools in aging research.

## Data Availability

Data from the VMAP cohort can be accessed freely following data use approval (www.vmacdata.org). The source code for the analysis conducted in this study is available on request.

## Bibliography

Ahmadi, K., Pereira, J. B., Westen, D. van, Pasternak, O., Zhang, F., Nilsson, M., Stomrud, E., Spotorno, N., & Hansson, O. (2024). Fixel-Based Analysis Reveals Tau-Related White Matter Changes in Early Stages of Alzheimer’s Disease. Journal of Neuroscience, 44(18). 10.1523/JNEUROSCI.0538-23.2024

Archer, D. B., Moore, E. E., Shashikumar, N., Dumitrescu, L., Pechman, K. R., Landman, B. A., Gifford, K. A., Jefferson, A. L., & Hohman, T. J. (2020). Free-water metrics in medial temporal lobe white matter tract projections relate to longitudinal cognitive decline. Neurobiol Aging, 94, 15–23. 10.1016/j.neurobiolaging.2020.05.001

Aung, W. Y., Mar, S., & Benzinger, T. L. (2013). Diffusion tensor MRI as a biomarker in axonal and myelin damage. Imaging Med, 5(5), 427–440. 10.2217/iim.13.49

Bartzokis, G., Cummings, J. L., Sultzer, D., Henderson, V. W., Nuechterlein, K. H., & Mintz, J. (2003). White Matter Structural Integrity in Healthy Aging Adults and Patients With Alzheimer Disease: A Magnetic Resonance Imaging Study. Archives of Neurology, 60(3), 393–398. 10.1001/archneur.60.3.393

Bartzokis, G., Sultzer, D., Lu, P. H., Nuechterlein, K. H., Mintz, J., & Cummings, J. L. (2004). Heterogeneous age-related breakdown of white matter structural integrity: Implications for cortical “disconnection” in aging and Alzheimer’s disease. Neurobiology of Aging, 25(7), 843–851. 10.1016/j.neurobiolaging.2003.09.005

Bender, A. R., Weissman, C., Scheel, N., Jung, Y., Damoiseaux, J. S., Peltier, S. J., & Hampstead, B. M. (2025). Multivariate analysis of white matter crossing fibers reveals correlations with aMCI and dementia diagnoses. Alzheimer’s & Dementia, 20(Suppl 2), e087463. 10.1002/alz.087463

Bendlin, B. B., Fitzgerald, M. E., Ries, M. L., Xu, G., Kastman, E. K., Thiel, B. W., Rowley, H. A., Lazar, M., Alexander, A. L., & Johnson, S. C. (2010). White Matter in Aging and Cognition: A Cross-sectional Study of Microstructure in Adults Aged Eighteen to Eighty-Three. Developmental Neuropsychology, 35(3), 257–277. 10.1080/87565641003696775

Benitez, A., Jensen, J. H., Falangola, M. F., Nietert, P. J., & Helpern, J. A. (2018). Modeling white matter tract integrity in aging with diffusional kurtosis imaging. Neurobiology of Aging, 70, 265–275. 10.1016/j.neurobiolaging.2018.07.006

Bhattarai, A., Maillard, P., Decarli, C., & Fan, A. (2023). Fixel based analysis of white matter alterations in Mild cognitive impairment and Alzheimer’s disease. Alzheimer’s & Dementia, 19(S16), e079454. 10.1002/alz.079454

Billaud, C. H. A., Yu, J., & Alzheimer’s Disease Neuroimaging Initiative. (2024). Fixel-based and tensor-derived white matter abnormalities in relation to memory impairment and neurocognitive disorders. GeroScience. 10.1007/s11357-024-01340-8

Braak, H., Del Tredici, K., Schultz, C., & Braak, E. (2000). Vulnerability of Select Neuronal Types to Alzheimer’s Disease. Annals of the New York Academy of Sciences, 924(1), 53–61. 10.1111/j.1749-6632.2000.tb05560.x

Brickman, A. M., Meier, I. B., Korgaonkar, M. S., Provenzano, F. A., Grieve, S. M., Siedlecki, K. L., Wasserman, B. T., Williams, L. M., & Zimmerman, M. E. (2012). Testing the white matter retrogenesis hypothesis of cognitive aging. Neurobiology of Aging, 33(8), 1699–1715. 10.1016/j.neurobiolaging.2011.06.001

Charlton, R. A., Barrick, T. R., McIntyre, D. J., Shen, Y., O’Sullivan, M., Howe, F. A., Clark, C. A., Morris, R. G., & Markus, H. S. (2006). White matter damage on diffusion tensor imaging correlates with age-related cognitive decline. Neurology, 66(2), 217–222. 10.1212/01.wnl.0000194256.15247.83

Cox, S. R., Ritchie, S. J., Tucker-Drob, E. M., Liewald, D. C., Hagenaars, S. P., Davies, G., Wardlaw, J. M., Gale, C. R., Bastin, M. E., & Deary, I. J. (2016). Ageing and brain white matter structure in 3,513 UK Biobank participants. Nature Communications, 7(1), Article 1. 10.1038/ncomms13629

Dhollander, T., Clemente, A., Singh, M., Boonstra, F., Civier, O., Duque, J. D., Egorova, N., Enticott, P., Fuelscher, I., Gajamange, S., Genc, S., Gottlieb, E., Hyde, C., Imms, P., Kelly, C., Kirkovski, M., Kolbe, S., Liang, X., Malhotra, A., … Caeyenberghs, K. (2021). Fixel-based Analysis of Diffusion MRI: Methods, Applications, Challenges and Opportunities. NeuroImage, 241, 118417. 10.1016/j.neuroimage.2021.118417

Douaud, G., Jbabdi, S., Behrens, T. E. J., Menke, R. A., Gass, A., Monsch, A. U., Rao, A., Whitcher, B., Kindlmann, G., Matthews, P. M., & Smith, S. (2011). DTI measures in crossing-fibre areas: Increased diffusion anisotropy reveals early white matter alteration in MCI and mild Alzheimer’s disease. NeuroImage, 55(3), 880–890. 10.1016/j.neuroimage.2010.12.008

Ferro, J. M., & Madureira, S. (2002). Age-related white matter changes and cognitive impairment. Journal of the Neurological Sciences, 203–204, 221–225. 10.1016/S0022-510X(02)00295-2

Figley, C. R., Uddin, M. N., Wong, K., Kornelsen, J., Puig, J., & Figley, T. D. (2022). Potential Pitfalls of Using Fractional Anisotropy, Axial Diffusivity, and Radial Diffusivity as Biomarkers of Cerebral White Matter Microstructure. Frontiers in Neuroscience, 15. 10.3389/fnins.2021.799576

Finkelstein, A., Faiyaz, A., Weber, M. T., Qiu, X., Uddin, M. N., Zhong, J., & Schifitto, G. (2021). Fixel-Based Analysis and Free Water Corrected DTI Evaluation of HIV-Associated Neurocognitive Disorders. Frontiers in Neurology, 12, 725059. 10.3389/fneur.2021.725059

Fjell, A. M., McEvoy, L., Holland, D., Dale, A. M., & Walhovd, K. B. (2014). What is normal in normal aging? Effects of Aging, Amyloid and Alzheimer’s Disease on the Cerebral Cortex and the Hippocampus. Progress in Neurobiology, 117, 20–40. 10.1016/j.pneurobio.2014.02.004

GarniercCrussard, A., Bougacha, S., Wirth, M., Dautricourt, S., Sherif, S., Landeau, B., Gonneaud, J., De Flores, R., de la Sayette, V., Vivien, D., KrolakcSalmon, P., & Chételat, G. (2022). White matter hyperintensity topography in Alzheimer’s disease and links to cognition. Alzheimer’s & Dementia, 18(3), 422–433. 10.1002/alz.12410

Gong, Z., Bilgel, M., Kiely, M., Triebswetter, C., Ferrucci, L., Resnick, S. M., Spencer, R. G., & Bouhrara, M. (2023). Lower myelin content is associated with more rapid cognitive decline among cognitively unimpaired individuals. Alzheimer’s & DementiaJ: The Journal of the Alzheimer’s Association, 19(7), 3098–3107. 10.1002/alz.12968

Haykal, S., Ćurčić-Blake, B., Jansonius, N. M., & Cornelissen, F. W. (2019). Fixel-Based Analysis of Visual Pathway White Matter in Primary Open-Angle Glaucoma. Investigative Ophthalmology & Visual Science, 60(12), 3803–3812. 10.1167/iovs.19-27447

Hua, K., Oishi, K., Zhang, J., Wakana, S., Yoshioka, T., Zhang, W., Akhter, K. D., Li, X., Huang, H., Jiang, H., van Zijl, P., & Mori, S. (2009). Mapping of Functional Areas in the Human Cortex Based on Connectivity through Association Fibers. Cerebral Cortex, 19(8), 1889–1895. 10.1093/cercor/bhn215

Huang, H., Fan, X., Weiner, M., Martin-Cook, K., Xiao, G., Davis, J., Devous, M., Rosenberg, R., & Diaz-Arrastia, R. (2012). Distinctive disruption patterns of white matter tracts in Alzheimer’s disease with full diffusion tensor characterization. Neurobiology of Aging, 33(9), 2029–2045. 10.1016/j.neurobiolaging.2011.06.027

Jeurissen, B., Leemans, A., Tournier, J., Jones, D. K., & Sijbers, J. (2012). Investigating the prevalence of complex fiber configurations in white matter tissue with diffusion magnetic resonance imaging. Human Brain Mapping, 34(11), 2747–2766. 10.1002/hbm.22099

Jeurissen, B., Tournier, J.-D., Dhollander, T., Connelly, A., & Sijbers, J. (2014). Multi-tissue constrained spherical deconvolution for improved analysis of multi-shell diffusion MRI data. NeuroImage, 103, 411–426. 10.1016/j.neuroimage.2014.07.061

Kennedy, K. M., & Raz, N. (2009). Aging white matter and cognition: Differential effects of regional variations in diffusion properties on memory, executive functions, and speed. Neuropsychologia, 47(3), 916–927. 10.1016/j.neuropsychologia.2009.01.001

Kirkovski, M., Fuelscher, I., Hyde, C., Donaldson, P. H., Ford, T. C., Rossell, S. L., Fitzgerald, P. B., & Enticott, P. G. (2020). Fixel Based Analysis Reveals Atypical White Matter Micro- and Macrostructure in Adults With Autism Spectrum Disorder: An Investigation of the Role of Biological Sex. Frontiers in Integrative Neuroscience, 14. 10.3389/fnint.2020.00040

Kochunov, P., Ramage, A. E., Lancaster, J. L., Robin, D. A., Narayana, S., Coyle, T., Royall, D. R., & Fox, P. (2009). Loss of cerebral white matter structural integrity tracks the gray matter metabolic decline in normal aging. NeuroImage, 45(1), 17–28. 10.1016/j.neuroimage.2008.11.010

Kuczynski, B., Targan, E., Madison, C., Weiner, M., Zhang, Y., Reed, B., Chui, HC., & Jagust, W. (2010). White matter integrity and cortical metabolic associations in aging and dementia. Alzheimer’s & DementiaJ: The Journal of the Alzheimer’s Association, 6(1), 54. 10.1016/j.jalz.2009.04.1228

Landman, B. A., Bogovic, J. A., Wan, H., ElShahaby, F. E. Z., Bazin, P.-L., & Prince, J. L. (2012). RESOLUTION OF CROSSING FIBERS WITH CONSTRAINED COMPRESSED SENSING USING DIFFUSION TENSOR MRI. Neuroimage, 59(3), 2175–2186. 10.1016/j.neuroimage.2011.10.011

Lee, S.-H., Coutu, J.-P., Wilkens, P., Yendiki, A., Rosas, H. D., & Salat, D. H. (2015). Tract-Based Analysis of White Matter Degeneration in Alzheimer’s Disease. Neuroscience, 301, 79–89. 10.1016/j.neuroscience.2015.05.049

Liu, H., Yang, Y., Xia, Y., Zhu, W., Leak, R. K., Wei, Z., Wang, J., & Hu, X. (2017). Aging of Cerebral White Matter. Ageing Research Reviews, 34, 64–76. 10.1016/j.arr.2016.11.006

Luo, X., Wang, S., Jiaerken, Y., Li, K., Zeng, Q., Zhang, R., Wang, C., Xu, X., Wu, D., Huang, P., & Zhang, M. (2021). Distinct fiber-specific white matter reductions pattern in early- and late-onset Alzheimer’s disease. Aging, 13(9), 12410–12430. 10.18632/aging.202702

Madden, D. J., Spaniol, J., Costello, M. C., Bucur, B., White, L. E., Cabeza, R., Davis, S. W., Dennis, N. A., Provenzale, J. M., & Huettel, S. A. (2009). Cerebral white matter integrity mediates adult age differences in cognitive performance. Journal of Cognitive Neuroscience, 21(2), 289–302. 10.1162/jocn.2009.21047

Mayo, C. D., Mazerolle, E. L., Ritchie, L., Fisk, J. D., & Gawryluk, J. R. (2017). Longitudinal changes in microstructural white matter metrics in Alzheimer’s disease. NeuroImage: Clinical, 13, 330–338. 10.1016/j.nicl.2016.12.012

Mito, R., Raffelt, D., Dhollander, T., Vaughan, D. N., Tournier, J.-D., Salvado, O., Brodtmann, A., Rowe, C. C., Villemagne, V. L., & Connelly, A. (2018). Fibre-specific white matter reductions in Alzheimer’s disease and mild cognitive impairment. Brain: A Journal of Neurology, 141(3), 888–902. 10.1093/brain/awx355

Moffett, J. R., Ross, B., Arun, P., Madhavarao, C. N., & Namboodiri, M. A. A. (2007). N-Acetylaspartate in the CNS: From Neurodiagnostics to Neurobiology. Progress in Neurobiology, 81(2), 89–131. 10.1016/j.pneurobio.2006.12.003

Nasrabady, S. E., Rizvi, B., Goldman, J. E., & Brickman, A. M. (2018). White matter changes in Alzheimer’s disease: A focus on myelin and oligodendrocytes. Acta Neuropathologica Communications, 6(1), 22. 10.1186/s40478-018-0515-3

Newman, B. T., Dhollander, T., Reynier, K. A., Panzer, M. B., & Druzgal, T. J. (2020). Test-retest reliability and long-term stability of 3-tissue constrained spherical deconvolution methods for analyzing diffusion MRI data. Magnetic Resonance in Medicine, 84(4), 2161–2173. 10.1002/mrm.28242

Nguyen, T.-T., Cheng, J.-S., Chen, Y.-L., Lin, Y.-C., Tsai, C.-C., Lu, C.-S., Weng, Y.-H., Wu, Y.-M., Hoang, N.-T., & Wang, J.-J. (2021). Fixel-Based Analysis of White Matter Degeneration in Patients With Progressive Supranuclear Palsy or Multiple System Atrophy, as Compared to Parkinson’s Disease. Frontiers in Aging Neuroscience, 13. 10.3389/fnagi.2021.625874

Oh, S. L., Chen, C.-M., Wu, Y.-R., Valdes Hernandez, M., Tsai, C.-C., Cheng, J.-S., Chen, Y.-L., Wu, Y.-M., Lin, Y.-C., & Wang, J.-J. (2021). Fixel-Based Analysis Effectively Identifies White Matter Tract Degeneration in Huntington’s Disease. Frontiers in Neuroscience, 15. 10.3389/fnins.2021.711651

O’Sullivan, M., Jones, D. K., Summers, P. E., Morris, R. G., Williams, S. C., & Markus, H. S. (2001). Evidence for cortical “disconnection” as a mechanism of age-related cognitive decline. Neurology, 57(4), 632–638. 10.1212/wnl.57.4.632

Papuć, E., & Rejdak, K. (2018). The role of myelin damage in Alzheimer’s disease pathology. Archives of Medical ScienceJ: AMS, 16(2), 345–351. 10.5114/aoms.2018.76863

Peter, C., Sathe, A., Shashikumar, N., Pechman, K. R., Workmeister, A. W., Jackson, T. B., Huo, Y., Mukherjee, S., Mez, J., Dumitrescu, L. C., Gifford, K. A., Bolton, C. J., Gaynor, L. S., Risacher, S. L., Beason-Held, L. L., An, Y., Arfanakis, K., Erus, G., Davatzikos, C., … Archer, D. B. (2025). White Matter Abnormalities and Cognition in Aging and Alzheimer Disease. 10.1001/jamaneurol.2025.1601

Petersen, M., Frey, B. M., Mayer, C., Kühn, S., Gallinat, J., Hanning, U., Fiehler, J., Borof, K., Jagodzinski, A., Gerloff, C., Thomalla, G., & Cheng, B. (2022). Fixel based analysis of white matter alterations in early stage cerebral small vessel disease. Scientific Reports, 12, 1581. 10.1038/s41598-022-05665-2

Raffelt, D. A., Smith, R. E., Ridgway, G. R., Tournier, J.-D., Vaughan, D. N., Rose, S., Henderson, R., & Connelly, A. (2015). Connectivity-based fixel enhancement: Whole-brain statistical analysis of diffusion MRI measures in the presence of crossing fibres. NeuroImage, 117, 40–55. 10.1016/j.neuroimage.2015.05.039

Raffelt, D. A., Tournier, J.-D., Smith, R. E., Vaughan, D. N., Jackson, G., Ridgway, G. R., & Connelly, A. (2017). Investigating white matter fibre density and morphology using fixel-based analysis. NeuroImage, 144(Pt A), 58–73. 10.1016/j.neuroimage.2016.09.029

Raffelt, D., Tournier, J.-D., Rose, S., Ridgway, G. R., Henderson, R., Crozier, S., Salvado, O., & Connelly, A. (2012). Apparent Fibre Density: A novel measure for the analysis of diffusion-weighted magnetic resonance images. NeuroImage, 59(4), 3976–3994. 10.1016/j.neuroimage.2011.10.045

Salvadores, N., Sanhueza, M., Manque, P., & Court, F. A. (2017). Axonal Degeneration during Aging and Its Functional Role in Neurodegenerative Disorders. Frontiers in Neuroscience, 11. 10.3389/fnins.2017.00451

Sánchez, S. M., Duarte-Abritta, B., Abulafia, C., De Pino, G., Bocaccio, H., Castro, M. N., Sevlever, G. E., Fonzo, G. A., Nemeroff, C. B., Gustafson, D. R., Guinjoan, S. M., & Villarreal, M. F. (2020). White matter fiber density abnormalities in cognitively normal adults at risk for late-onset Alzheimer’s disease. Journal of Psychiatric Research, 122, 79–87. 10.1016/j.jpsychires.2019.12.019

Schilling, K. G., Tax, C. M. W., Rheault, F., Landman, B., Anderson, A., Descoteaux, M., & Petit, L. (2021). Prevalence of white matter pathways coming into a single diffusion MRI voxel orientation: The bottleneck issue in tractography (p. 2021.06.22.449454). bioRxiv. 10.1101/2021.06.22.449454

Stone, D. B., Ryman, S. G., Hartman, A. P., Wertz, C. J., Vakhtin, A. A., & Alzheimer’s Disease Neuroimaging Initiative. (2021). Specific White Matter Tracts and Diffusion Properties Predict Conversion From Mild Cognitive Impairment to Alzheimer’s Disease. Frontiers in Aging Neuroscience, 13. 10.3389/fnagi.2021.711579

Tinney, E. M., Warren, A. E. L., Ai, M., Morris, T. P., O’Brien, A., Odom, H., Sutton, B. P., Jain, S., Kang, C., Huang, H., Wan, L., Oberlin, L., Burns, J. M., Vidoni, E. D., McAuley, E., Kramer, A. F., Erickson, K. I., & Hillman, C. H. (2024). Understanding Cognitive Aging Through White Matter: A Fixel-Based Analysis. Human Brain Mapping, 45(18), e70121. 10.1002/hbm.70121

Voineskos, A. N., Rajji, T. K., Lobaugh, N. J., Miranda, D., Shenton, M. E., Kennedy, J. L., Pollock, B. G., & Mulsant, B. H. (2012). Age-related decline in white matter tract integrity and cognitive performance: A DTI tractography and structural equation modeling study. Neurobiology of Aging, 33(1), 21–34. 10.1016/j.neurobiolaging.2010.02.009

Wen, Q., Mustafi, S. M., Li, J., Risacher, S. L., Tallman, E., Brown, S. A., West, J. D., Harezlak, J., Farlow, M. R., Unverzagt, F. W., Gao, S., Apostolova, L. G., Saykin, A. J., & Wu, Y.-C. (2019). White matter alterations in early-stage Alzheimer’s disease: A tract-specific study. *Alzheimer’s & Dementia*J*: Diagnosis*, Assessment & Disease Monitoring, 11, 576–587. 10.1016/j.dadm.2019.06.003

Wiegell, M. R., Larsson, H. B. W., & Wedeen, V. J. (2000). Fiber Crossing in Human Brain Depicted with Diffusion Tensor MR Imaging. Radiology, 217(3), 897–903. 10.1148/radiology.217.3.r00nv43897

